# Pulmonary hypertension misdiagnosis due to preventable errors in echocardiography and right heart catheterization

**DOI:** 10.1101/2024.06.19.24309207

**Authors:** Alexandra Saunders, Kevin R Bainey, Rhea Varughese, Evangelos D Michelakis

## Abstract

**Objectives:** We hypothesized that preventable human errors in performance and reporting of transthoracic echocardiograms (TTEs) and right RHCs are common and may lead to misdiagnosis of pulmonary hypertension (PH) subgroups.

**Background:** PH is a common disease, however PH subgroups have vastly different mortality and treatment. This is particularly the case for pulmonary arterial hypertension (PAH) versus PH secondary to heart failure with preserved ejection fraction (HFpEF). TTE) and RHC are needed to differentiate these two diseases. Diagnosis requires specific cut-offs for mean pulmonary artery pressure (mPAP) and pulmonary artery wedge pressure (PAWP), which can only be measured by RHC. However, TTE first identifies PH, triggering referral to specialized PH centres.

**Methods:** We re-analyzed TTEs and RHCs of 252 PH program referrals over 5 years. We also compared the inferred diagnosis from the original reports to the diagnosis made after error correction.

**Results:** We identified numerous preventable errors in the performance and reporting of both tests, and subsequently there was a poor correlation between the parameters measured by both tests. The referral TTE reports missed or overcalled PH in 44 patients. The RHC, mostly by PAWP mistakes, led to misdiagnosis in 41 patients.

**Conclusion:** TTE errors may delay referrals, while RHC errors may lead to misdiagnosis and applying wrong therapies to patients. As PAH therapies are extremely expensive, this also impacts the health care system. Primary care physicians need to be on alert for such errors and referral centres need to promote quality improvement programs to mitigate these errors.

## Introduction

Pulmonary hypertension (PH) is a disease with high morbidity and mortality, with the prognosis varying among its subgroups. Even with therapy, pulmonary arterial hypertension (PAH; Group 1) mortality is high with a 59% 5-year survival – worse than many metastatic cancers(1,2). Therapies are expensive with significant side effects and are unable to reverse the disease. Late diagnosis also worsens mortality by >20%(3). While PAH is rare, Group 2 PH (secondary to left ventricular dysfunction) is very common(4). The most difficult Group 2 subtype to differentiate from Group 1 PAH is PH due to heart failure with preserved ejection fraction (HFpEF)(4), which has significant treatment and prognosis differences from Group 1. Recently, promising classes of drugs have been shown to be effective in HFpEF(5). There are also approved therapies for the other groups of PH(6). Thus, the type of PH needs to be diagnosed accurately and early, with referral to a specialized PH center. These centers are available in most states, where the management of the biggest driver of mortality, the development of right ventricular failure, can be monitored along with the optimal timing for lung transplantation, the ultimate treatment for advanced PAH. Misdiagnosing PH types will not only impact the patient, but also the health care system, since the cost of treating PAH exceeds $200,000 per year (7,8).

Two tests are paramount in the workup for PH: transthoracic echocardiography (TTE) and right heart catheterization (RHC). TTE is usually the first indication that the patient has PH and leads to the referral to a specialized PH center, where RHC is performed. RHC is the only test that can reliably separate Group 1 (where PAWP needs to be ≤15 mmHg) from Group 2 (where PAWP needs to be >15 mmHg(6,9)), as TTE cannot measure PAWP. Additionally, TTE can only estimate systolic PA pressure (sPAP), while RHC can directly measure it and dPAP (diastolic PA pressure) to give mPAP (mean PA pressure). This and the cardiac output (CO) are required for a PH label (mPAP > 20 mmHg(6)) and for the calculation of pulmonary vascular resistance (PVR=mPAP-PAWP/CO), as PVR >3 WU is also needed to diagnose PH(6,9). In addition, PVR is required to understand whether a patient responds to vasodilators, since all current PAH therapies are vasodilators and effective therapy requires a larger drop in PVR than systemic vascular resistance (SVR).

TTE uses many biophysical assumptions to estimate sPAP and RV function. Slight deviation from best practice can lead to unreliable results. Similarly, RHC requires meticulous attention to detail of hemodynamic tracings and the art of careful RHC varies from center to center. Thus, the possibility of test-based mistakes leading to PH misdiagnosis (or potential for missed diagnosis) is not to be ignored. Importantly, while cardiologists may be more aware of the vulnerability of TTE and RHC to mistakes and review the actual tests themselves, most physicians taking care of PH patients are non-cardiologists with less knowledge of Doppler signals on TTE and hemodynamic variability during RHC. Across the 17 PH clinics currently in Canada, 34 physicians are respirologists and 8 are cardiologists(10). The implication of this is that there are less physicians who are experienced with TTE and RHC interpreting these results. Referrals from other physicians are dependent on the TTE and RHC reports for timely referral or application of guideline-directed medical therapy. We aimed to a) assess the frequency of mistakes included in the performance and reports of TTE and RHC, and b) determine how often those mistakes could lead to misdiagnosis or missed diagnosis, in the setting of a large Canadian university hospital referral center for PH.

## Methods

### Study Design

At the University of Alberta PH program, patients undergo RHC prior to initiating PAH therapy. We re-reviewed the original data from all TTE’s that prompted a referral to our PH center over the last 5 years, along with the initial RHC that followed the referring TTE, and recorded errors in the performance and the reporting. All the TTE’s and RHC’s were reviewed by two cardiologists that were different from the interpreting cardiologist. The quality of the over-read of the TTE and RHC data was assessed by comparing 20 TTEs and RHCs between the two readers, which showed a 95% agreement. Our study received appropriate approvals from the University of Alberta Ethics Board.

We determined whether the data reported by the TTE correlated with the data reported by the RHC (the gold standard), particularly for parameters measured by both tests (e.g., right ventricular systolic pressure; RVSP). We also determined whether the data describing RV function (tricuspid annular plane systolic excursion; TAPSE) and TTE RV size or cardiac output and PA pulsatility index (PAPi) in the RHC correlate with WHO (World Health Organization) functional class assessment and the 6-minute walk tests recorded in the same time period as the TTE and RHC.

After recording the errors in the interpretation and reporting, we also compared the diagnosis inferred by the tests reports, to the diagnosis after correction of the TTE and RHC errors. We focused on the parameters that need to be included in a TTE report or in a RHC report to determine the presence and severity of pulmonary hypertension, the response to acute vasodilators (inhaled nitric oxide), and the response to chronic PAH therapies (relative decrease in PVR over SVR), as shown in **Table 1**. For TTE, this included right atrial pressure estimation, tricuspid regurgitation (TR) velocity, TR grade, RVSP, TAPSE, RV dilation grade, and if there were other potential causes for high left atrial (LA) pressure. Each TTE was reviewed to see if poor TR jet quality was mentioned, if the TR was not checked in multiple views, if the TAPSE was recorded, if RV hypertrophy was present, if RV dilatation was quantified, and if the right atrial pressure (RAP) was estimated appropriately using the inferior vena cava diameter and distensibility. Each RHC was reviewed for calibration quality (e.g., if RAP reported was lower than −3 mmHg). They were also reviewed for whether respiratory variation for PAWP was recorded and if it was recorded correctly (as the gold standard is to record end-expiratory values). The PAWP also should be the average of 3 different beats and the left ventricular end-diastolic pressure (LVEDP) should not be different from the PAWP (as without mitral stenosis or pulmonary vein stenosis, these should be the same) or greater than the dPAP (which should not be physiologically possible as a gradient is needed, and clearly suggests error).

**Table 1.** Data collected for TTE and RHC studies and their rationale.

| **Parameter Reported on TTE** | **Rationale** |
| --- | --- |
| University vs community site | For comparison of quality of studies. |
| RVSP (ie: sPAP) | Typically, if >40 mmHg, PH or elevation of right-sided pressures is listed in the conclusion of the report. |
| Indication of whether TR velocity envelope is adequate (checking of envelope or if checked for in 3 views) | If poor TR jet quality is present, it should raise suspicion for potentially unreliable estimates of sPAP. |
| RAP (based on inferior vena cava distensibility and collapsibility) | This is needed to calculate sPAP, as opposed to an arbitrary value of 5 - 15 mmHg used in some centers. |
| RV free wall thickness | Suggestive of chronic and/or compensated PH. |
| RV size (RV dilation grade) | Suggestive of likelihood of decompensated RV function. |
| RV contractility (TAPSE) | Indicates RV decompensation if <2.0 cm. |
| **Parameters Reported on RHC** | **Rationale** |
| RAP | For comparison to TTE RAP and to determine if zeroing was appropriate (as this value should not be <-3 mmHg). |
| sPAP, dPAP, mPAP | For comparison to TTE RVSP. Also, to diagnose PH and to confirm quality of PAWP (as dPAP should be >PAWP). |
| PAWP waveform quality (O<sub>2</sub> saturation to confirm wedge position, average of 3 cardiac cycles, and end-expiratory recording) | Validates the accuracy of the measurement. If not done correctly, this should raise suspicion that the value of PAWP, and therefore PVR, may not be accurate. |
| LVEDP | Validates the accuracy of the reported PAWP. |
| CO (thermodilution or modified Fick) | The accuracy and clinical application of CO varies between thermodilution and modified Fick methods, and thus so does the calculation of PVR. |
| PVR | Used in the diagnosis of PH and vulnerable to errors in both PAWP and CO. |

### Study Population

260 consecutive patients referred to our PH program with an TTE, subsequently undergoing RHC. Of these, 6 patients were excluded due to severe mitral stenosis or severe pulmonic stenosis, so that this did not cloud the accuracy of comparing PAWP to LVEDP or TTE RVSP compared to RHC sPAP. Two patients were also excluded for severe LV systolic failure, i.e., EF<40%, to ensure that patients with elevated PAWPs reflected HFpEF rather than HFrEF, giving a final sample size of 252. TTEs were performed on various echo machines and utilizing different software programs depending on the site, while RHCs all were interpreted using Philips IntelliVue X3® program at the University hospital.

Baseline characteristics of the population studied include 57% female, mean age of 57, mean sPAP of 56±1.6 mmHg, mean PAWP of 14±0.6 mmHg, WHO functional class 3±0.05, 6 min walk distance 343±16 m. Additionally, 27% of the patients had already died at the time of analysis of the data spanning a 5-year period, representing that this is a sick group of PH patients and typical for populations assessed in referral centers. Original inferred diagnosis was PAH for 111 and HFpEF for 78 patients.

### Statistical Analysis

To assess the relationship between the TTE parameters and the gold standard RHC measures, as well as to assess the reported RHC PAWP values with the gold standards of end-expiration measurement and LVEDP measurement, we reported Pearson correlation coefficients. We also reported Pearson correlation coefficients for CO vs WHO functional class and 6-minute-walk distance.

A regression analysis will show whether these values correlate, but not if they agree or the degree of agreement. For this reason, we also utilized a Bland Altman analysis to look at the agreement between the mean values of the two tests, showing the limits and potential bias of one variable compared to the gold standard variable. The Bland Altman analysis plots the difference for each paired value of the 2 tests over the mean difference of all the paired values.

## Results

### 1. TTE

Several errors were identified in the performance of the TTEs and their interpretations (**Table 2**).

**Table 2.** Frequency of potential measurement errors (left) and number of misses/misclassifications of PH attributed to these errors.

| **Parameters on TTE** | **Number of patients (%)** | **Type of misdiagnosis from TTE errors**<br>• PH missed: 9 (4)<br>• PH overcalled: 35 (14)<br>-----<br>**Type of misdiagnosis from RHC errors in PAWP**<br>• True PAH labelled as HFpEF: 21 (8)<br>• True HFpEF labelled as PAH: 20 (8) |
| --- | --- | --- |
| Poor quality of TR envelope not reported | 61 (34) |  |
| TR not checked in 3 views | 21 (12) |  |
| RAP not appropriately based on inferior vena cava size and collapsibility | 23 (9) |  |
| RV free wall thickness not reported | 245 (97) |  |
| RV size not reported | 9 (4) |  |
| RV contractility not reported (and without rationale, like poor image quality) | 41 (16) |  |
| **Parameters on RHC** |  |  |
| PAWP position not confirmed using PAWP O<sub>2</sub> saturation | 252 (100) |  |
| PAWP not reported at end-expiratory stage (when respiratory variation was recorded) | 60 (24) |  |
| PAWP recorded during a breath hold | 81 (32) |  |
| PAWP not the average of 3 cardiac cycles | 26 (10) |  |
| PAWP > 5 mmHg difference from LVEDP | 48 (38) |  |
| PAWP > dPAP | 25 (10) |  |
| CO method not done using thermodilution | 217 (86) |  |

The RVSP_TTE_ vs sPAP_RHC_ was only moderately correlated (r = 0.62) with slight improvement if looking at RHCs done less than two days from the TTE (r = 0.76), compared to r = 0.60 when > 2 weeks (**Figure 1**). The mean deviation of RVSP_TTE_ from sPAP_RHC_ (gold standard) was 18 mmHg (± 1) and Bland-Altman analysis showed 95% of the differences ranged from −40 to +50 mmHg. This wide degree of spread was fairly similar between the University hospital and the community sites (both with r = 0.6) as was the degree of differences on the Bland-Altman analysis (Figure 1A), suggesting that this difference was not center-specific. To explain this variation, when reviewing the TTEs for an indication of TR jet quality, we found that many studies confidently reported a value for RVSP when the TR jet quality was poor (34% of studies; **Table 2**). The TR jet was also not always checked in at least 3 views (12% of studies).

**Figure 1:**
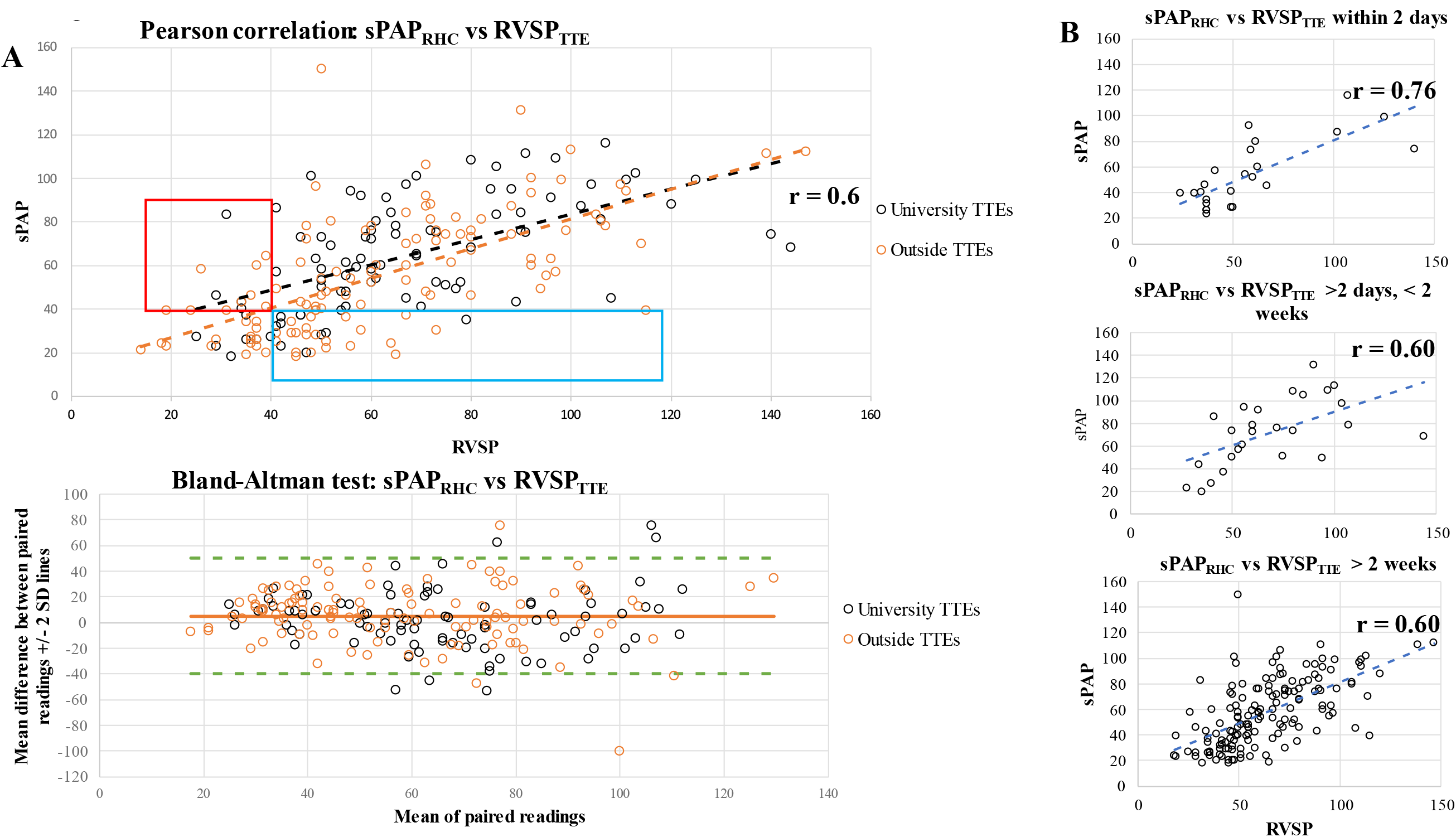
All sPAP_RHC_ values compared to patient-matched RVSP_TTE_ values, with black dots marking studies performed at the University echocardiography lab, and orange dots at outside center echocardiography labs. All RHC were done at the University site. **1A: Top**: A Pearson correlation is shown. The values only correlated moderately and with the same r value of 0.6, whether they were University or community TTEs. **Bottom**: A Bland-Altman analysis of the paired TTE/RHC values shows a wide range of disagreement between the two tests, higher than what is considered clinically “acceptable”. The red box shown patients where TTE would have missed the diagnosis of PH (typically reported if RVSP is >40 mmHg) since the same patients had higher values of sPAP in the RHC. The blue box represents patients where TTE stated that there was PH, when there was no PH in the RHC (sPAP<40 mmHg). **1B**: The same Pearson correlation is shown in subgroups where the 2 tests were done within 2 days from each other, between 2 days and 2 weeks, and >2 weeks.

RV function was not always quantified. Moreover, 16% of studies did not report a TAPSE value (or mention that image quality was too poor to evaluate TAPSE). TAPSE was weakly correlated with sPAP (r = −0.27), unsurprisingly, as many other factors compromise RV contractility over and above increased afterload. RV thickness was usually not commented on (97% of studies did not mention RV thickness). The degree of RV dilatation was qualitatively reported in most of the TTEs (96%), with 48% of studies reporting some degree of RV dilation.

Since the PAPi (sPAP-dPAP)/RAP) is a RHC value often cited as a surrogate of RV function, we compared PAPi with RV function and size on TTE. There was no correlation between TAPSE and PAPi (r = 0.05, **Figure 2A**) or between the degree of RV dilation and PAPi (r = −0.08, **Figure 2B**). PAPi also had no correlation with 6-minute walk distance (r = 0.08) or WHO-functional class (r = −0.13; not shown).

**Figure 2:**
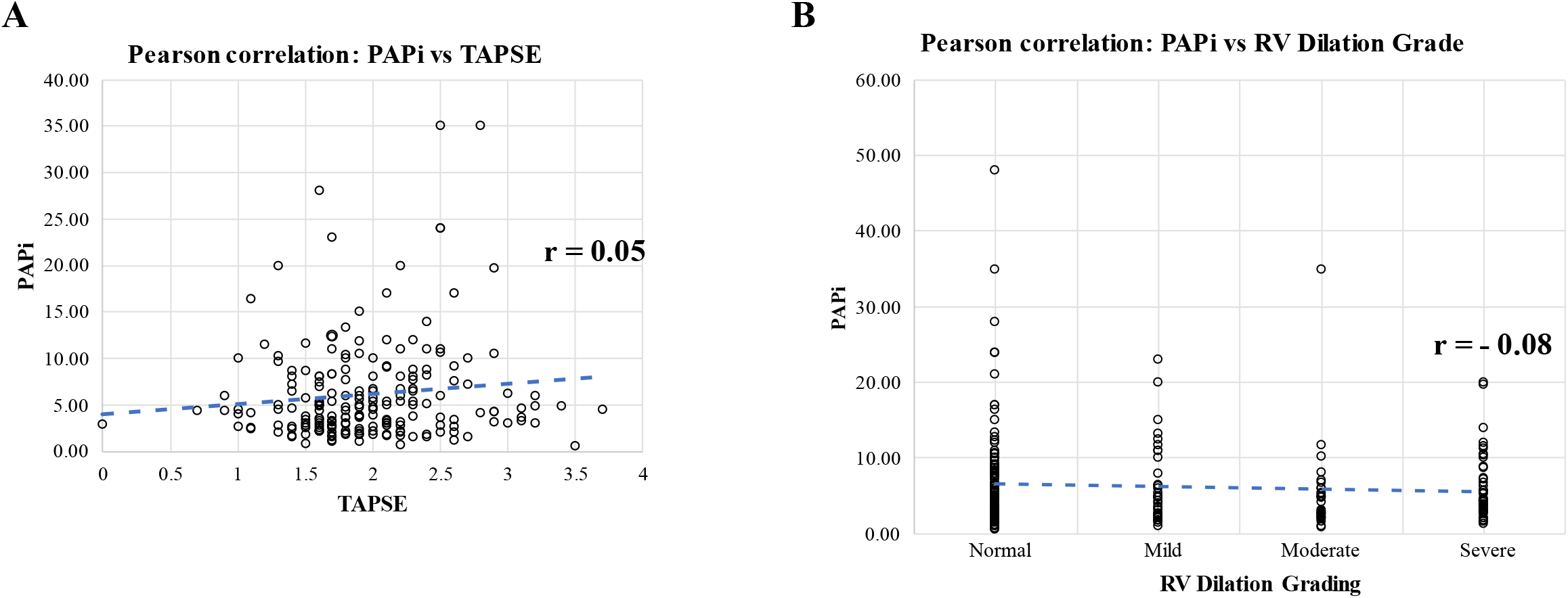
Scatterplot of RHC-based calculated PAPi vs TTE TAPSE, showing no correlation **(A)**. There is also no correlation between PAPi and RV size at end-diastole reported on the TTE **(B**).

RAP estimation was done incorrectly in 10% of TTEs. When comparing the RAP estimation on TTE to RHC, the overall correlation was poor. As this value is very dependent on loading conditions and volume status, the data was grouped into those who had TTE and RHC within two days, within 2 days to 2 weeks, and greater than two weeks of each other; this still did not improve the correlation by a significant amount (r value of 0.5, 0.6, 0.4, respectively).

Overall, there were several instances where measurements on TTE could have led to the wrong conclusion for a patient diagnosis. Typically, a mention of PH should be included in the TTE report if the estimated RVSP is higher than 40 mmHg. As shown in the red box in **Figure 1A** and in **Table 2**, 9 patients would have been missed as having PH if a RHC had not also been done (i.e., the TTE reported a normal RVSP, when sPAP was truly elevated on RHC and in some cases higher than 60 or even 80 mmHg). Importantly, although these TTEs all had other features of PH (RV dilation, reduced RV function), “possible PH” or a statement that RV pressures could be underestimated was not mentioned in the reports. This is important because the falsely normal RVSP report could prevent an ordering physician from sending a referral to a PH center. Several patients were also labelled as having PH on TTE, when the RHC found pressures to be normal (35 patients, **Table 2**), which has the potential for misdiagnosis, since there was no therapeutic intervention between the TTE and the RHC.

### 2. RHC

Several errors were identified in the performance of the RHCs (**Table 2**).

When reviewing respiratory variation of the PAWP and the final reported value, 60 studies did not properly report the end-expiratory PAWP (e.g. reporting on inspiration or the mean between inspiration and expiration), while 81 studies utilized a single PAWP value (as opposed to 3 averaged values) with no specification of the respiratory cycle at all. Of those done incorrectly, 3 patients had their PAWP reported as >15 mmHg when it was truly low, which could have missed a diagnosis of PAH over HFpEF. Nine patients had their PAWP reported as <15 mmHg when it was truly higher, which could have given them an incorrect diagnosis of PAH rather than HFpEF. There could have been more of these misclassifications, as it is unclear if these patients were truly in end-expiration for the value reported (since 81 studies did not record respiratory variation).

There were 120 studies that included a left-heart catheterization, so we were able to compare the LVEDP directly to the PAWP reported (as this value should be nearly equivalent to the LVEDP in the absence of mitral stenosis, which was excluded from our study). The Pearson correlation for LVEDP vs PAWP was only 0.50 and the Bland-Altman analysis showed that 95% of the difference between LVEDP and PAWP fell between −11.3 and +14.9 mmHg (**Figure 3a**). With the reported PAWP there were 37/124 (29.8%) studies that had a difference greater than 5 mmHg (the acceptable level of difference). This relationship slightly improved if the LVEDP was compared to the corrected true end-expiratory PAWP (r = 0.64, 95% agreement between −9.9 to +14.3; **Figure 3b**). When comparing the LVEDP to the PAWP in those studies that did not record any respiratory variation compared to those that did, the relationship was much worse. With no respiratory variation recorded, LVEDP vs PAWP had an r = 0.27 and Bland-Altman analysis 95% agreement between −20.8 to +12.9 mmHg. With no respiratory variation recorded, 16/42 (38%) patients had >5 mmHg variation between PAWP and LVEDP. However, when respiratory variation was recorded (and interpreted properly, in retrospect) the r increased to 0.67, and the Bland-Altman analysis 95% agreement was between −9.6 to +8.7 mmHg. In that case, fewer patients (11/52, 21.1%) had >5 mmHg deviation (**Figure 3c & d**). As outlined by the red and blue boxes in Figure 3, several patients could have been misclassified if relying on the PAWP alone: the red boxes showing those patients who had a PAWP reported as >15 mmHg, but the LVEDP was actually <15 mmHg (18 patients) and the blue boxes showing those who had a PAWP reported as <15 mmHg, but the LVEDP was actually >15 mmHg (11 patients). Thus, in addition to the 12 patients that were misdiagnosed based on our re-analysis of the PAWP waveforms alone, 29 more patients were misdiagnosed based on the discrepancy between PAWP and LVEDP, for a total of 41 patients (**Table 2**).

**Figure 3:**
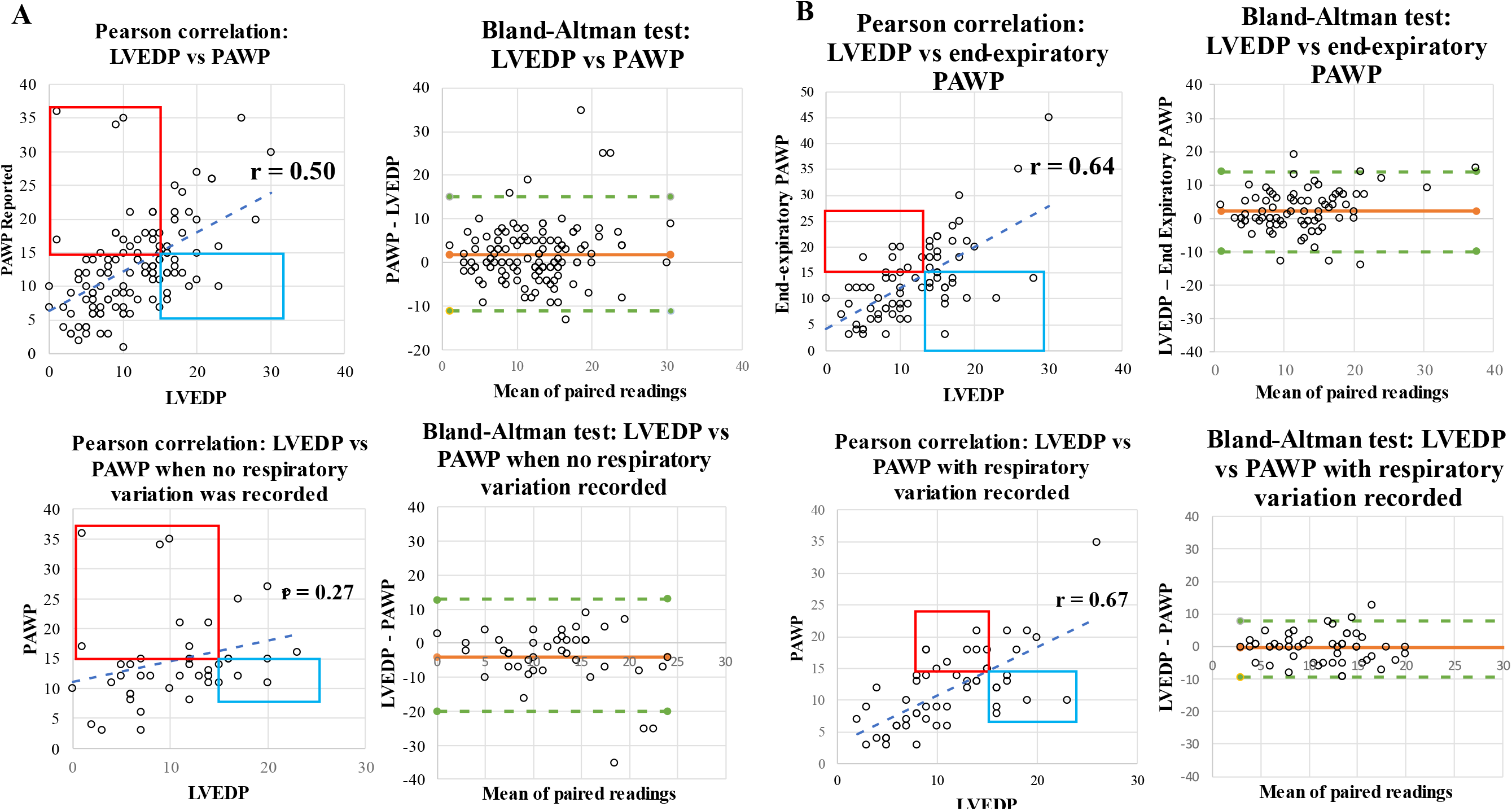
Pearson correlations and Bland-Altman analyses for comparisons of the LVEDP against various forms of the PAWP measurements: **A:** LVEDP versus PAWP reported in the same study of the same patient. **B:** LVEDP versus the corrected PAWP during our re-analysis of the RHC recorded pressure tracings. Comparing A to B shows that the PAWP traces were not interpreted correctly, as our reanalysis increased the r value toward LVEDP (i.e., the gold standard for the accuracy of the PAWP in the absence of mitral stenosis, which was excluded in our studies) **C:** LVEDP versus the reported PAWP when no respiratory variation was stated in the report and the PAWP was recorded during a breath hold. **D:** LVEDP versus the reported PAWP when the report clearly included traces showing respiratory variation of the PAWP value. Comparing C to D shows that the physicians sensitized to correctly recording and reporting PAWP were much closer to the gold standard value (i.e., LVEDP). Red boxes indicate patients where the PAWP was reported as high (suggesting HFpEF) while the LVEDP was <15 mmHg, thus missing a diagnosis of PAH. Blue boxes indicate patients where the PAWP was reported as low, suggesting PAH, while the true LVEDP was >15 mmHg, giving an incorrect label of PAH and missing PH secondary to HFpEF.

Despite these examples of inaccuracies in how the PAWP was being measured, no PAWP saturations were performed to confirm placement, and there were only three studies in which they reportedly could not obtain a wedge. There were also 25 studies where the PAWP measurement did not make physiological sense, as it was a greater value than the dPAP, certainly pointing to an error.

CO measurements were done in all RHC studies. The majority (86%; 217 studies) were done using the modified Fick’s method alone, while 9 studies used thermodilution alone, and 26 used both modified Fick’s method and thermodilution. In the small subset of studies that used both, we were able to compare CO measurements, and they did not correlate well, with an r value of 0.44 for Fick vs thermodilution and Bland-Altman analysis showed that 95% agreement range from −2.2 to +2.6 (**Figure 4**).

**Figure 4.**
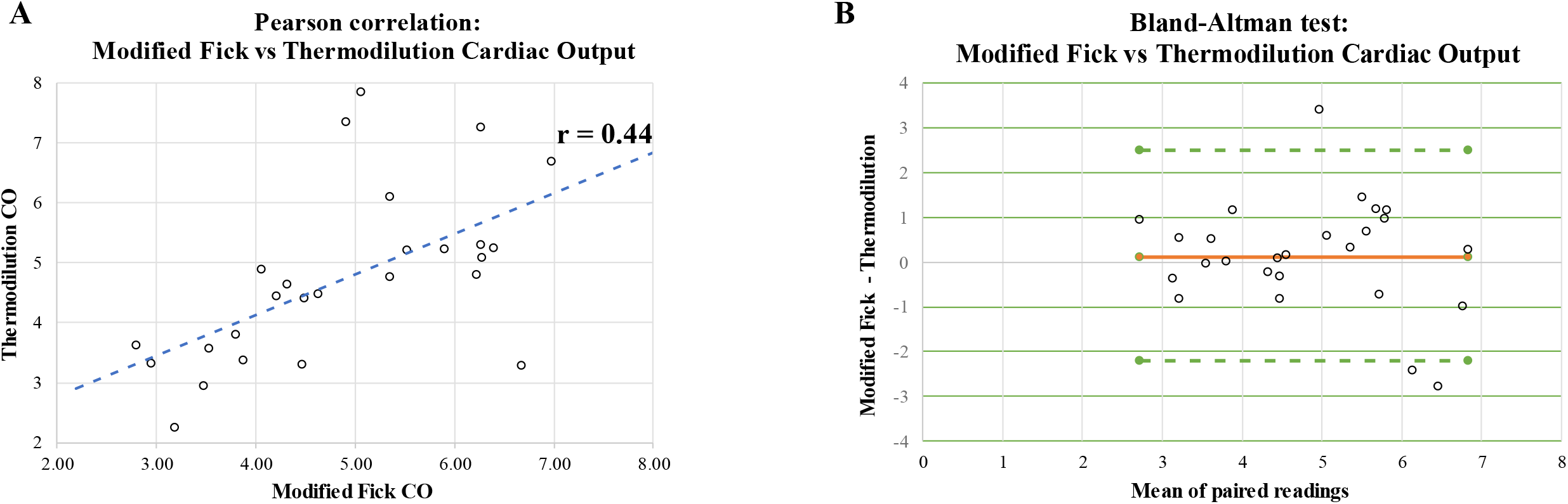
Pearson correlation and Bland-Altman analysis between the modified Fick cardiac output vs thermodilution cardiac output performed in the same study of the same patient show poor correlation and wide degree of disagreement.

There were several instances where the PVR value changed from >3 WU (in keeping with PH) to <3 WU, depending on which measure of CO was used. When correcting for PAWP (e.g. by using the correct end-expiratory PAWP to calculate PVR using a Fick CO) the PVR went from <3 WU to >3 Wood units 33 times and from >3 WU to <3 WU 8 times. When using the correct end-expiratory PAWP and a thermodilution CO, 3 out of 35 studies changed PVR category. Therefore, the accuracy of the PAWP and the method of CO measurement can both have a large impact on the PVR calculations and thus have clinical implications.

Due to restrictions from the COVID-19 pandemic that took place during our study period, there were 185 clinical assessments closely synchronized with TTE and RHC assessments that recorded WHO functional status and only 61 that recorded 6-minute walking distance. The 6-minute walking distance and WHO functional class had an expected negative correlation to each other: as the WHO functional class increased from I to IV, the 6-minute walking distance worsened, with an r value of −0.74. WHO functional class and 6-minute walking distance both did not correlate with Fick CO values but there was a moderate correlation with thermodilution CO values, with an r value of −0.46 and 0.45 respectively (**Figure 5**). There was also no correlation between CO and TAPSE or grade of RV dilation (not shown).

**Figure 5:**
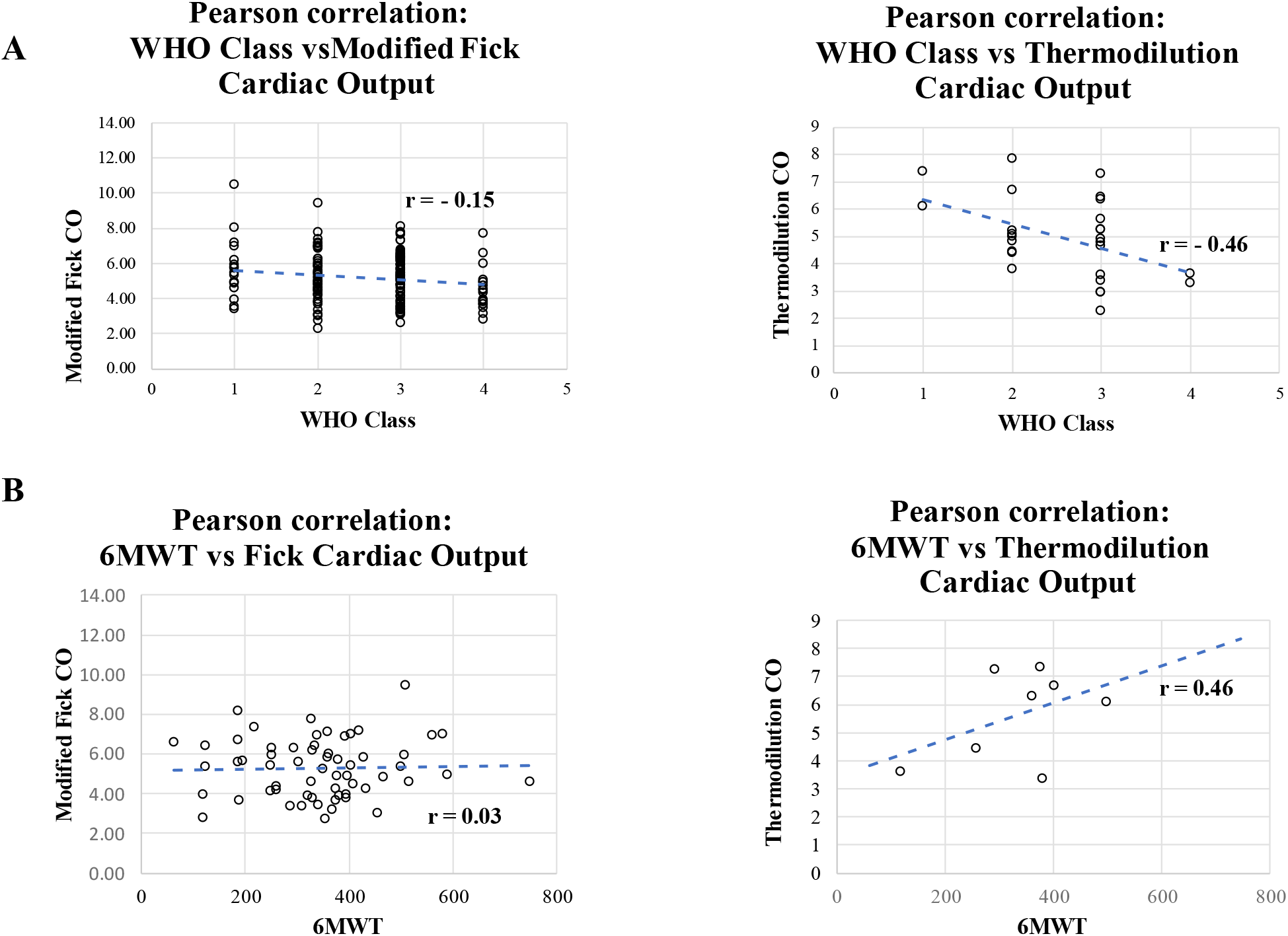
Pearson correlations of WHO functional class (**A**) and 6-minute walk distance (**B**) assessed close to the time of RHC to either modified Fick-measured (**left**) or thermodilution-measured cardiac output (**right**). The modified Fick cardiac output did not correlate at all with these two clinical parameters that is expected to relate with and unfortunately was the method used in the majority of RHCs.

## Discussion

We report that preventable errors can commonly occur during the performance and reporting of TTEs and RHCs, even in specialized referral centers, with a significant impact in the care of PH patients as well as the health care system. Overall, these errors affected diagnosis in 34 % of the referred PH patients (**Table 2**). The implications of these errors include:

**1)** Delays in the identification of possible PH at the primary care level, which in turn may cause delays in the referral of patients to specialized PH centers. The delay of PAH diagnosis is known to adversely affect the survival of patients and the optimal management of RV failure(3).
**2)** Misdiagnosis of PAH versus PH due to HFpEF. Those two conditions have significant differences in prognosis and treatment. At the specialist level, this is particularly relevant after the recent introduction of therapies that can be of significant benefit in HFpEF and prevent RV failure(5). As **Table 2** shows, over 16% of patients can suffer from misdiagnosis.
**3)** At the level of specialists that manage PH patients (including cardiologists, respirologists, rheumatologists, and hematologists), the mistakes in PVR can lead to misinterpretation of the response of these patients to PAH-therapies, since they are all vasodilators aiming for a preferential decrease in PVR over SVR. This may lead to wrong timing and application of guideline-directed medical therapies.
**4)** Considering the momentous costs of PAH therapies, these errors can have a significant impact on the health care system in addition to the application of the wrong medications (and thus their side effects) in the wrong patient population.

The majority of the TTE errors could be improved by robust TR jet interrogation and a careful statement of both the quality of the RVSP estimation and the description of the RV size and function. The condition of the RV (e.g., hypertrophy, dilatation), needs to also be emphasized if abnormal, since it may indicate PH, even if the estimated RVSP is less than 40. These could have important implications on whether a primary care physician will order with subsequent testing, referral to another specialist, or referral to a specialized PH center.

Similarly, most RHC errors can be fixed by the careful confirmation of appropriate PAWP position (with an O_2_ saturation check) and recording PAWP at the end-expiratory position for at least 3 cardiac cycles. Another conclusion from our work is that the modified Fick method of calculating CO does not appear to correlate with the functional status of the patients or the TTE-measured function of the RV. Reasons for this have been well described(11), resting with the several levels of estimation in the equation (CO = VO_2_/Ca-Cv). VO_2_ is estimated by using body surface area, multiplied by a constant. In our center, a constant value of 133 is used (differing from most studies, which often use the Dehmer formula of 125 x body surface area(12)). The 133 value is a proprietary default setting by the company supplying our catheterization program (Philips) and has not been studied or validated in published literature to our knowledge. Studies on the accuracy of these estimation methods show that other formulas may have somewhat better accuracy (like the LaFarge & Miettinen formula(13)) so one area for potential improvement is a change to the VO_2_ estimation formula. Despite these known limitations, the vast majority of RHCs performed used the modified Fick method, presumably as it is faster to acquire.

A limitation of this study is that the comparison of TTE with RHC should ideally be attempted when the two studies are performed simultaneously, perhaps as part of a research protocol. However, this is not realistic in clinical practice. In fact, a strength of our study is that it reflects routine clinical practice, increasing its relevance to both non-specialists and specialists dealing with PH. Another strength of this study is that it is the first, to our knowledge, to assess both the quality of the TTE and RHC simultaneously and their impact on PH management.

Our results should alert echocardiography and cardiac catheterization laboratory staff as well. Although we focused on studies of patients referred to our center with PH as a possible diagnosis, these same errors would apply to all the patients undergoing these tests, potentially significantly increasing the impact of these errors, extending it to non-PH populations.

For the primary care physician ordering the initial TTE, as well for the non-cardiologist PH specialist, it is important to realize the vulnerabilities of TTE. If a patient has symptoms of right-heart failure, like peripheral edema, ascites, or hepatosplenomegaly with decreased functional capacity, the threshold for referral to a PH center should be low.

For the PH expert, it is important to ask the right questions when referring a patient for RHC and to emphasize why the accuracy of those values will be important. If the clinical suspicion is high and the quality of TTE and RHC suboptimal, testing may need to be repeated. For the cardiologists it is important to be reminded that although RHC is often considered a simple task (as compared to a high-risk intervention) mistakes in measurements as simple as recording a PAWP are common and can make a dramatic difference for a patient.

Finally, for the health care system at large, this should trigger conduction of quality improvement projects in echocardiography and cardiac catheterization laboratories. Thankfully, there are studies showing that these errors can be remedied through robust quality improvement programs. One study at an academic hospital in Germany showed that with coaching and education, RHC sPAP to TTE RVSP obtained an r value of 0.87(14). Another study of two large academic hospitals in Delaware, USA showed that a teaching intervention improved their correlation between several TTE and RHC parameters significantly. For example, RVSP_TTE_ vs RVSP_RHC_ had r values improve from 0.30 to 0.77 and TTE reports of diastolic dysfunction vs LVEDP had r values improve from 0.09 to 0.62(15). In another study, simply changing from measuring “the chin” instead of “the beard” for over-gained continuous wave tricuspid regurgitation Doppler signals significantly improved correlation of TTE with RHC(16). One study comparing PAWP measures with and without doing PAWP saturations found that only 50% of initial PAWP measurements were truly occlusive, and that when wedging was repeated until a true PAWP saturation was obtained, the average PAWP was often at least 5 mmHg lower, resulting in reclassification of PH group in 10% of those patients(17). With repetition and reinforcement, any suggested changes can become the standard of care and these improvements could be applicable to many centers which may have encounter similar errors, as this problem certainly exists outside our institution.

## Data Availability

Available upon request

## Acknowledgements

None

## Abbreviations

CO: Cardiac output
HFpEF: Heart failure with preserved ejection fraction
LVEDP: Left ventricular end-diastolic pressure
PAPi: Pulmonary artery pulsatility index
PAWP: Pulmonary artery wedge pressure
PH/PAH: Pulmonary hypertension/pulmonary arterial hypertension
PVR: Pulmonary vascular resistance
RAP: Right atrial pressure
RHC: Right heart catheterization RV Right ventricle
RVSP: Right ventricular systolic pressure
sPAP, dPAP, mPAP: systolic, diastolic, and mean pulmonary artery pressure
SVR: systemic vascular resistance
TR: Tricuspid regurgitation
TTE: Transthoracic echocardiogram
WHO: World Health Organization

